# Merging Adaptive Designs with Dynamic Infectious Disease Models Allows Faster and more Accurate Diagnostic Test Accuracy Studies in the Case of an Epidemic

**DOI:** 10.1101/2025.11.25.25340962

**Authors:** Denise Köster, Madhav Chaturvedi, Nicole Rübsamen, Mahnaz Badpa, André Karch, Antonia Zapf

## Abstract

**Background:** During epidemics with emerging infections, diagnostic tests directly inform model-based decision-making and thereby shape infection control strategies. However, diagnostic accuracy studies (DTA) assessing the validity of these tests must be conducted under severe time and data constraints. We investigated whether the integration of adaptive designs and of epidemic spread modelling for prevalence prediction can accelerate DTA studies during epidemics with emerging infections without compromising statistical validity.

**Methods:** We compared three designs in a large-scale simulation study using a COVID-19 use case: a fixed design; a standard adaptive design with unblinded interim analysis enabling early stopping or sample size adaptation; and an adaptive design additionally integrating a prevalence projection model to inform sample size re-estimation. Data-generating mechanisms were based on infectious disease models and realistic recruitment constraints. As decision rules we used in one simulation line WHO criteria for DTA studies for COVID-19 and in the other one more liberal performance thresholds. Across 1,440 factorial scenarios (5,000 replications each), we evaluated study duration, sample size requirements, statistical power as well as bias in estimates.

**Results:** Both adaptive designs enabled substantial operational gains. For the WHO thresholds, early stopping (for futility or infeasibility) occurred in 80% of adaptive simulations; early efficacy stops were rare. Under more liberal thresholds, early termination was less frequent, leading to more studies reaching final analysis. Required sample sizes under WHO criteria frequently exceeded 10,000 participants, making fixed designs practically infeasible. Adaptive designs identified infeasible scenarios early and avoided continuation. Under liberal thresholds, recalculated sample sizes in adaptive designs closely tracked theoretical needs up to the upper quartile, in contrast to fixed designs mirroring the low power commonly observed in real-world pandemic studies. Overall, adaptive designs shortened study duration when stopping early and prevented continuation of unpromising trials.

**Discussion:** Adaptive designs in DTA studies during epidemics with emerging pathogens improve feasibility by preventing unrealistic recruitment targets and enabling early abandonment of non-viable scenarios. When realistic performance thresholds are used, adaptive re-estimation produces sample sizes more aligned with statistical requirements without systematic operational penalties. These findings support the adoption of adaptive approaches in confirmatory DTA studies for emerging infections as a pragmatic response to time pressure and uncertainty.

## 1 Introduction

During an epidemic or pandemic, early and reliable diagnoses of infection are essential not only to determine the disease status of an individual, but also to enable primary epidemiological studies to establish values for important epidemiological parameters. For emerging infections, new diagnostic tests have to be developed during the early phases of the pandemic or epidemic, and their accuracy needs to be evaluated within a tight time frame.

The SARS-CoV-2 pandemic highlighted the essential role of diagnostic tests in infection control. They enabled the rapid identification of infections and played a significant part in monitoring disease spread and implementing containment measures.

In 2020 and 2021, the World Health Organization (WHO) [1] published comprehensive guidelines for development and usage of diagnostic tests for SARS-CoV-2. The guidance document [2] defined acceptable and desired limits for sensitivity (≥ 80%; ≥ 90%) and specificity (≥ 97%; ≥ 99%) for Antigen-detecting rapid diagnostic tests (Ag-RDTs). In the same year, a Cochrane review on rapid, point-of-care (POC) antigen and molecular-based tests for diagnosing SARS-CoV-2 [3] demonstrated that the accuracy of available diagnostic tests was much lower, especially sensitivity (on average 73% in symptomatic and 55% in asymptomatic individuals). The diagnostic test accuracy studies included in the review had large methodological heterogeneities with respect to study duration, sample sizes, and other aspects.

Lack in quality of diagnostic accuracy studies for emerging infections is often justified by the urgency in which the tests need to be developed. However, even minor deviations in the estimation of diagnostic accuracy might have detrimental effects on population-level decision-making.

Chaturvedi et al. [4, 5] highlighted how inaccurate assumptions about the accuracy of antibody tests affect the parametrisation and results of infectious disease models, which rely heavily on precise assumptions to make accurate predictions about the course of an epidemic.

In the following, we investigate the use of adaptive designs to speed up diagnostic test accuracy (DTA) studies without endangering their validity. We investigate modifications including early stopping for efficacy or futility, as well as incorporating infectious disease modelling into sample size re-estimations in DTA studies to enable better estimation of the target condition, which may change rapidly during an epidemic or pandemic.

## 2 Methods

### 2.1 General

During the early stages of an epidemic or a pandemic caused by an emerging pathogen, there is large uncertainty surrounding the epidemiological characteristics of the disease, as well as the true prevalence of the disease in the population. This makes it difficult to parameterise infectious disease models, as well as estimate sample sizes for DTA studies. We propose linking diagnostic test accuracy studies with infection spread models in an adaptive design to address both these issues. An unblinded interim analysis can be conducted to directly estimate parameters for the infection spread model (e.g. proportion of infection that is asymptomatic), or to provide initial estimates for sensitivity and specificity which, in conjunction with primary epidemiological studies, can be used to inform model parameterisation. In turn, the infectious disease model can be used to provide estimates of future prevalence of the target condition, which can then be used to re-estimate the sample size of the DTA study (see Figure 1).

**Figure 1:**
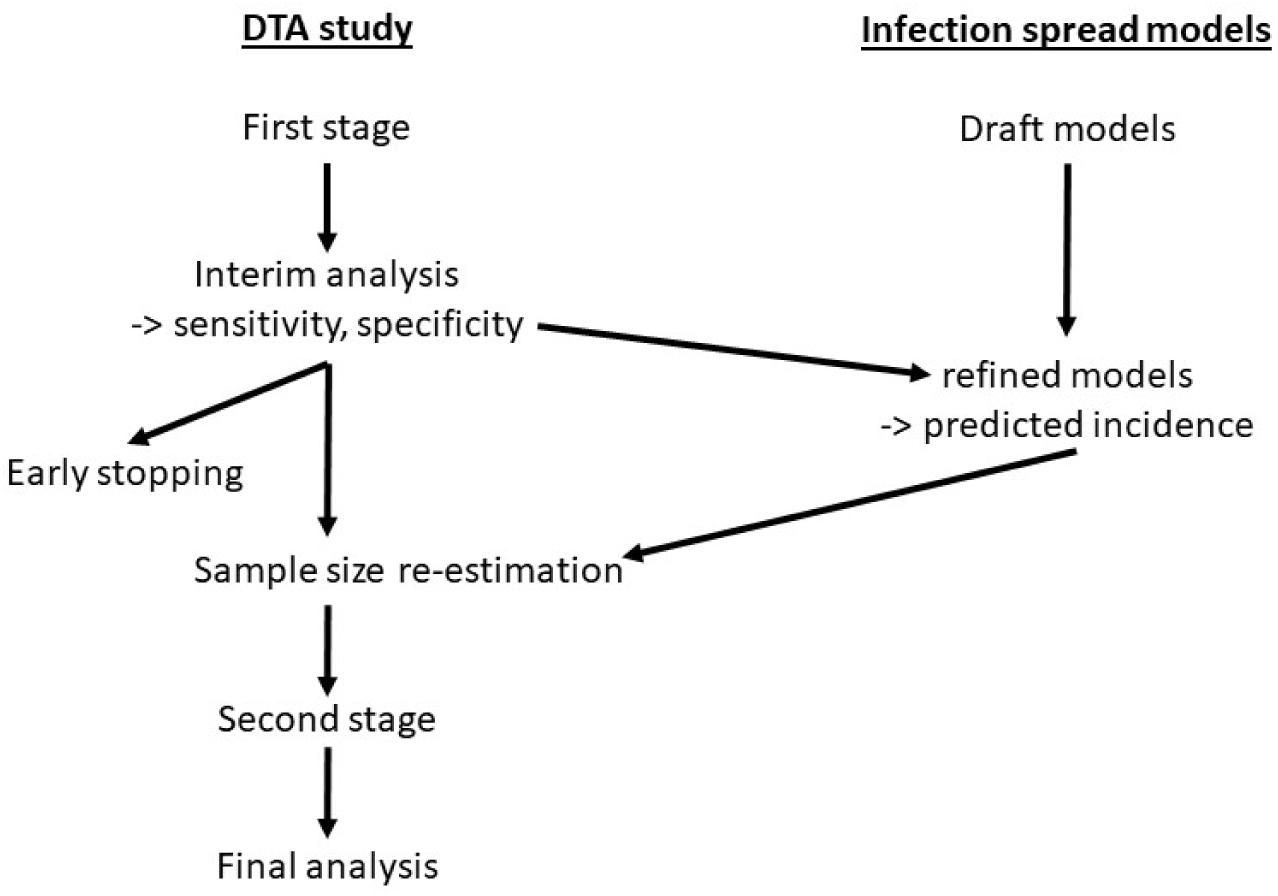
Visualisation of the overall study concept

The COVID-19 pandemic serves as a use case for this methodological framework, due to the extensive data availability and urgent need for accurate diagnostic tests during its early phases. Simulation scenarios were based on the Cochrane review of Ag-RDTs [3] and we took WHO’s published acceptance limits for SARS-CoV2 POC tests [2] as benchmark criteria for test performance in our simulations.

### 2.2 Hypotheses, sample size calculation and analyses

We investigated the single test design, where both the index test and the reference standard (which defines the true target condition) are administered to all participants. According to the recommendation of the WHO guidelines, the accuracy of the index test is measured by sensitivity (se) and specificity (sp), defined as the probability for a true positive result given the target condition’s presence (se) and a true negative result given its absence (sp). The individual hypotheses regarding the co-primary endpoints sensitivity and specificity in the DTA study are as follows [6]:

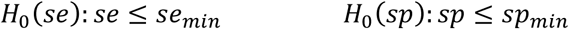

with *se_min_* and *sp_min_* as pre-defined minimum sensitivity and specificity. Linking the two hypotheses with an intersection union test results in the following global null hypothesis:

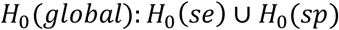

which requires both null hypotheses to be rejected for the global null to be rejected. the intersection union principle controls the overall type I error rate at the nominal level without requiring multiplicity adjustment. However, the power for each individual test must be increased to maintain sufficient overall power.

The sample size calculations are performed separately for each subgroup (those with and without the target condition) using the method proposed by Obuchowski [7]:

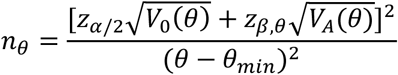

where *θ* is sensitivity or specificity, 𝜃*_min_* is corresponding minimum value, and 𝜃_A_ is the expected value under the alternative hypothesis, and V_0_(𝜃) and V_A_(𝜃) are variances under the null and the alternative hypothesis. The quantile 𝑧_𝛼/2_ refers to the quantile of the standard normal distribution for the given type-one error 𝛼, while 𝑧_𝛽,𝜃_ refers to the quantile for 1 minus the power for the respective endpoint. Furthermore, the ratio between the sample size of the subgroups should be representative for the subsequent intended use. To meet the overall target power, this study followed the approach of Stark and Zapf [8], which selects power for both endpoints to exceed a minimum while approaching the target.

For the analysis, we used logit confidence intervals, which are range-preserving and achieve the theoretical power for the sample size calculation used (see Stark and Zapf [8]):

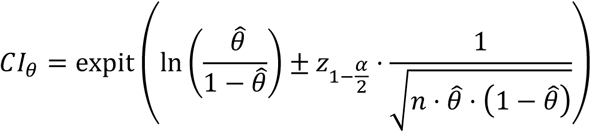

with 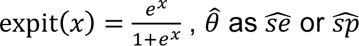, and 𝑛 corresponding to the number of individuals with or without target condition.

Furthermore, one-sample binomial tests were performed:

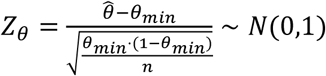

The individual hypotheses were rejected if the respective p-values are below the given type-one error and the global hypothesis was rejected if this condition is met for sensitivity and specificity.

### 2.2 Simulation Study

We conducted a comprehensive simulation study using the ADEMP structure (Aim, Data-generating mechanism, Estimand, Methods, Performance measures), in accordance with recommendations by Morris [9] and Burton [10].

#### Aim

We aimed to evaluate and compare the performance of three DTA study designs in the context of an evolving epidemic, including Fixed Design (FD): Standard design with no interim adaptations, Adaptive Design without modelling (ADs): Interim analysis and sample size re-estimation based on observed data only, and Adaptive Design with model-informed projections (ADp): Interim analysis and sample size re-estimation incorporating prevalence forecasts.

#### Data-generating mechanism

We simulated the spread of infection in a virtual population, using a stochastic compartmental model, described in Chaturvedi et al. [4] as the “true disease model”. Using the estimated prevalence from this simulation, we generated the data for the reference standard as binary test results and for the index test under the alternative hypothesis according to the given sensitivities and specificities. Furthermore, we selected minimum sensitivity and specificity as either *se_min_* = 80% and *sp_min_* = 97%, according to the defined WHO limits, or, more liberally, *se_min_* = 75% and *sp_min_* = 90%. The true values used for data generation ranged from 82.5% to 92.5% for sensitivity and from 97.5% to 99% for specificity.

Since no prior knowledge is available for sample size planning in the event of an emerging infection, we did not initially calculate the sample size here, but determined it based on information available from previous epidemics. In particular, initial sample sizes were derived from the first, second, and third quartiles of sample sizes in the Cochrane review in order to reflect realistic DTA study scales during the COVID-19 pandemic [3]. To capturing variability in study feasibility, recruitment rates corresponded to quartiles of enrolment rates from the same review. Moreover, we aligned study start dates (20, 40, 100 days post-pandemic onset) with early, peak, and declining epidemic phases, respectively, based on simulated incidence curves.

This factorial design yields 1,440 scenarios (2 thresholds × 5 sensitivities × 4 specificities × 3 initial sample sizes × 4 recruitment rates × 3 start dates), as shown in Table 1

**Table 1.** Simulated scenarios.

| Factor | $(se_{min}, sp_{min})$ | $se_A$ | $sp_A$ | Initial $N$ | Patients per day | Days since beginning of the pandemic | Overall |
| --- | --- | --- | --- | --- | --- | --- | --- |
| Values | (80%, 97%),<br>(75%, 90%) | 82.5%<br>to<br>92.5%<br>by<br>2.5%<br>steps | 97.5%<br>to<br>99%<br>by<br>0.5%<br>steps | 150,<br>328,<br>760 | 8,16,35,<br>100 | 20, 40, 100<br>days | Complete<br>factorial |
| Number of scenarios | 2 | 5 | 4 | 3 | 4 | 3 | 1,440 |

#### Estimand

The primary estimand was the statistical power regarding sensitivity and specificity estimates. Secondary estimands covered sample size and study duration.

#### Methods

We compared the adaptive design outlined in Section 2.1 and shown schematically in Figure 1 with a fixed design (FD) and with an adaptive design in which the variability in disease prevalence is not taken into account.

- The FD served as the control scenario and was implemented without any adaptive components. For each simulation run, the trial proceeded until the pre-specified total sample size was reached. All hypothesis testing and interval estimation was performed at the end of the study. No interim analyses were conducted, and the design did not allow for early stopping or sample size modifications. Sensitivity and specificity were assessed using two-sided binomial tests at the 5% significance level (see Section 2.2).
- In the standard adaptive design (ADs) without consideration of the dynamics of the pandemic, an unblinded interim analysis was conducted once 50% of the initial sample size had been recruited (Köster et al., 2026, under review). The interim analysis involved conducting hypothesis tests for both sensitivity and specificity using the combination testing approach proposed by Bauer and Köhne [11] in conjunction with the conditional error function from Proschan and Hunsberger [12]. The framework enabled sample size re-estimation while controlling the type I error rate.

In the interim analysis, two early stopping criteria were defined: 1) 𝛼𝛼_0_ for futility: If either interim p-value exceeded a predefined futility boundary (pre-specified, set to 0.8), the trial was terminated early due to futility; 2) 𝛼_1_ for efficacy: If both p-values of the first stage are below the efficacy boundary 𝛼_1_, the study can be stopped early for efficacy. In all other cases the sample size is recalculated and the study continues with the second stage. Sample size re-estimation was conducted based on the interim estimates of sensitivity, specificity, and observed disease prevalence with the assumption that prevalence remained constant for the remainder of the trial. After recruiting the remaining required sample size, the two p-values from the second stage were calculated and compared with the respective conditional error function 𝐴(𝑠_1,𝜃_). If both p-values are below the thresholds, the global null hypothesis could be rejected. The design is illustrated in Figure 2. Due to the great time pressure in a pandemic, we imposed an additional restriction for successful transition to the second stage—that the calculated sample size must be achieved in practice within one year (since the start of the pandemic).

- The adaptive design with predicted prevalence (ADp) differs from the ADs approach described above in that it takes into account the changing prevalence of the target condition by using an infection spread model to obtain estimates of the expected prevalence over the recruitment period. The mean prevalence over this period is then used for sample size re-estimation as described in the ADs approach. The model used for this purpose in these simulations is a deterministic compartmental model, described in detail as the “disease projection model” in Chaturvedi et al. [4]. The structure of this model corresponds to the “true disease model” used to generate the data i.e. in the simulation, it represents a “well-specified” model. In addition, all parameters except the proportion of asymptomatic infection were the same as the true disease model. The proportion of asymptomatic infection was estimated in the interim analysis of the DTA study as the ratio of participants displaying symptoms who tested positive using the reference test to all participants who tested positive with the reference test. The modelled course of the incidence over one year is shown for one example scenario in Figure 3. The challenge in taking this variable incidence into account when re-estimating the sample size is that the mean prevalence depends on the required recruitment period, which depends on the sample size, which in turn depends on the prevalence. The solution was an iterative procedure in which the available recruitment period is extended day by day, the mean prevalence for this period is calculated and then the corresponding sample size is calculated. Using the pre-specified number of patients that can be recruited per day, the required recruitment time for this sample size can then be inferred. As soon as the available and the required recruitment period are approximately the same (tolerance threshold 10%), the corresponding sample size is selected. The decision criteria for an early stopping are exactly the same as for ADs and the decision rules at the end of the possible second stage are also the same.

**Figure 2:**
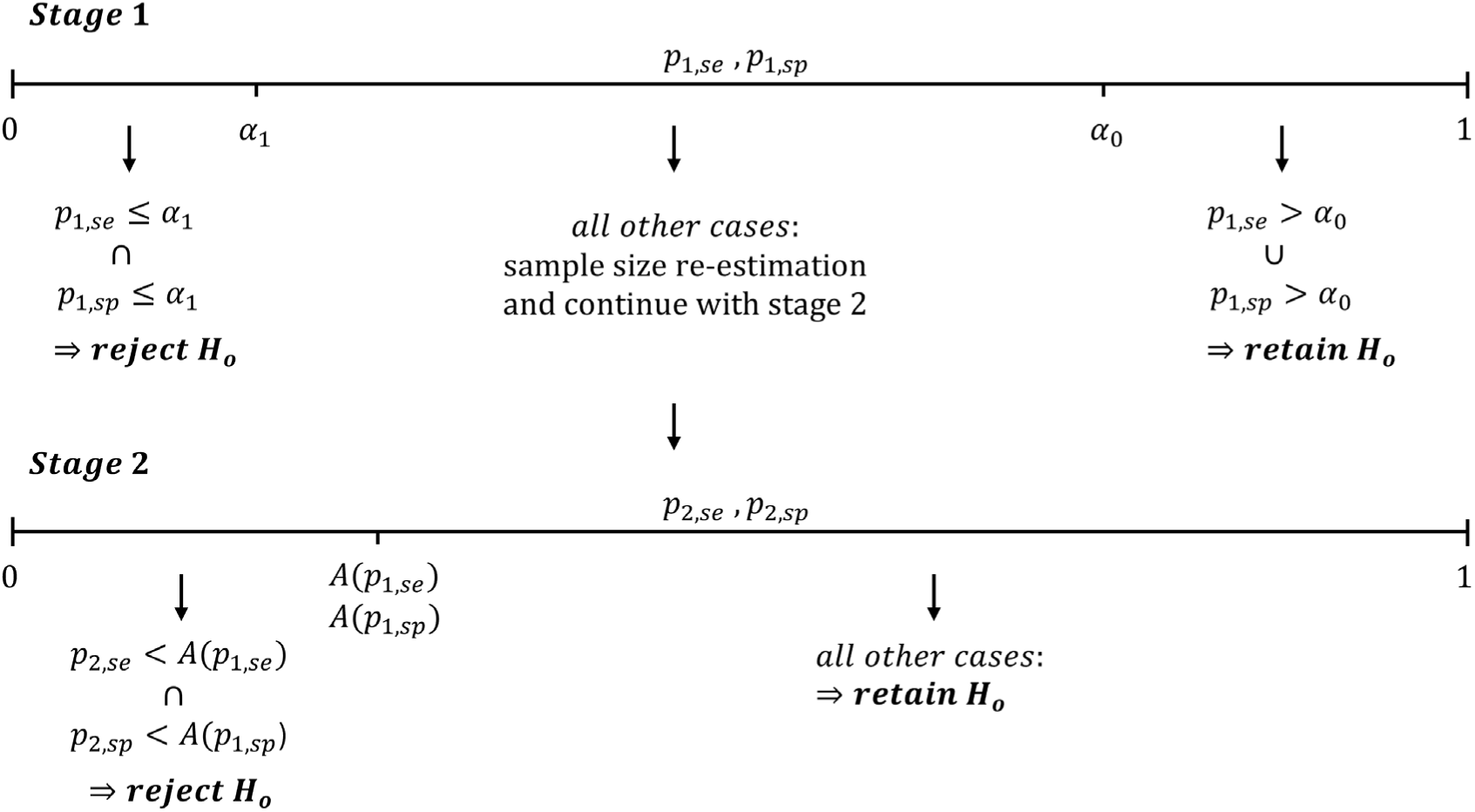
Illustration of the adaptive design with unblinded interim analysis and sample size re-estimation.

**Figure 3:**
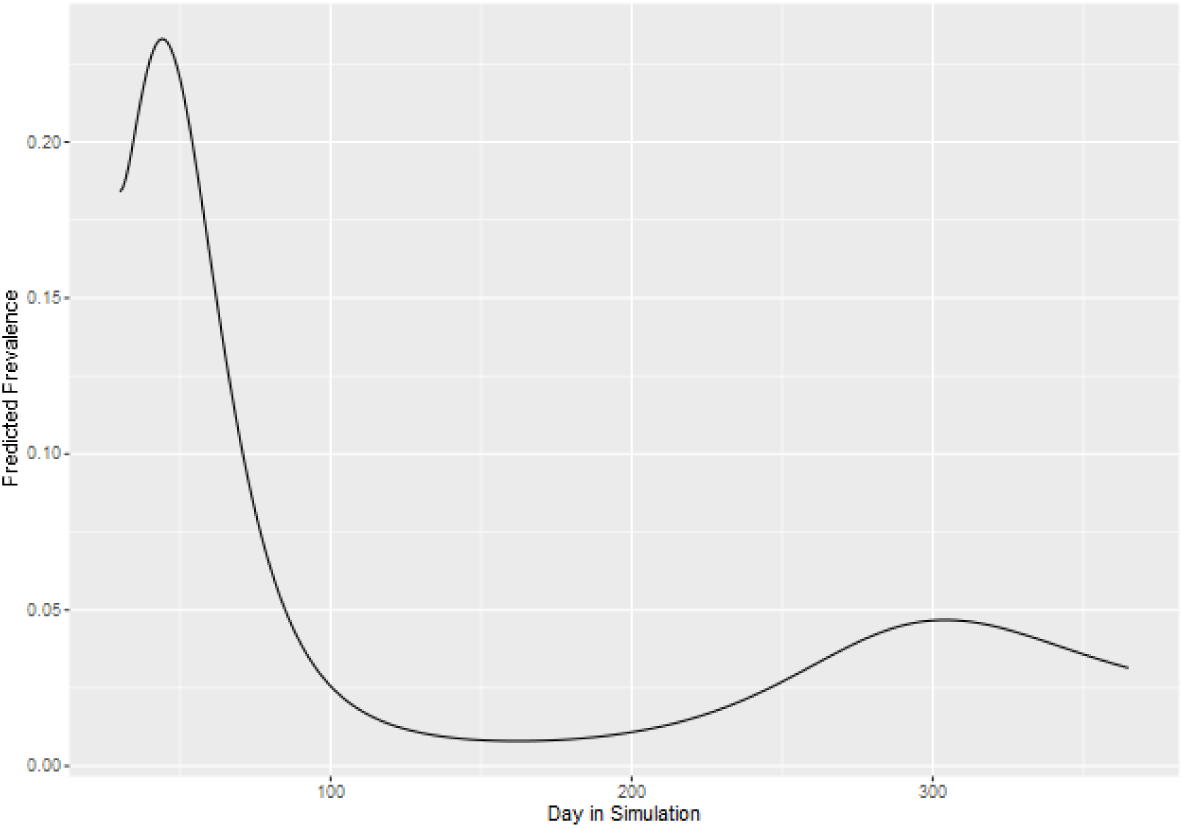
Modelled course of the incidence over one year for one example scenario.

#### Performance measures

The fixed design and the two adaptive designs were compared regarding the statistical power, efficiency by sample size, and relative bias. We conducted the simulation study in parallel on a high-performance-cluster running under Debian GNU/Linux 12 (bookworm) using R version 4.2.3 (2024-12-05). Each scenario was seeded with a unique random number, ensuring both reproducibility and independence of parallel computation. We evaluated each of the 1,440 scenarios with 5,000 simulation runs. The primary outcome of interest was the overall power over all simulation runs *n_sim_*, calculated as follows:

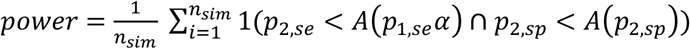

A further outcome parameter was the percentage bias:

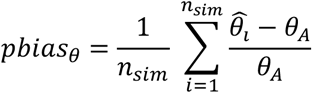

**Figure 4:**
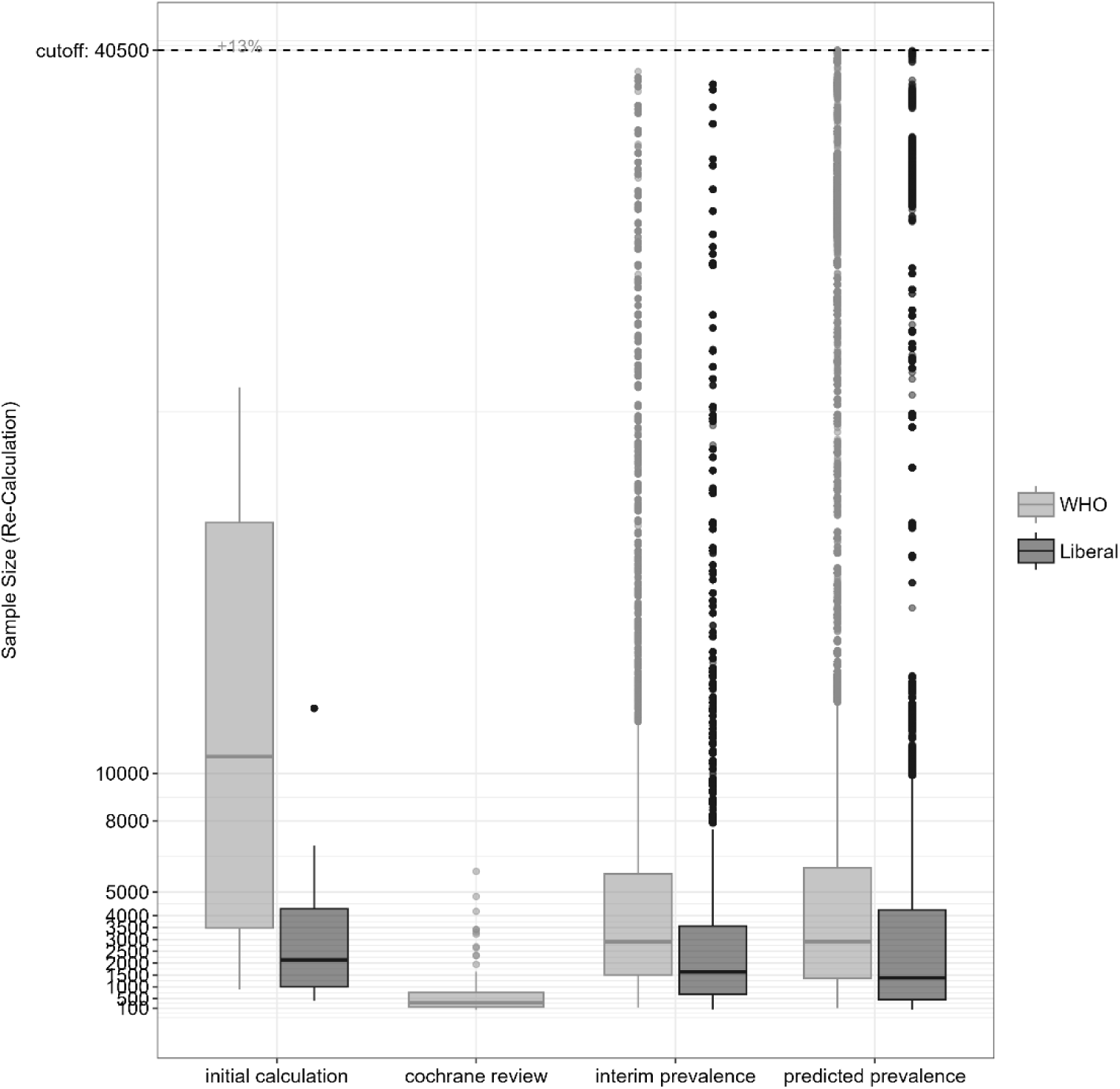
Theoretically needed sample size versus real sample sizes from the Cochrane review versus recalculated sample sizes of the adaptive designs.

## 4 Results

The key findings of the simulation study are summarized in Table 2, and are presented separately according to the accuracy thresholds applied and the type of study design used.

**Table 2:** Summary of simulation results by design and diagnostic threshold (WHO criteria and more liberal criteria) across 540 simulated scenarios with 5,000 replications each.

|  |  |  | Adaptive design |  | Fixed design (FD) |
| --- | --- | --- | --- | --- | --- |
|  |  |  | ADs | ADp |  |
| WHO criteria<br>$se_{min} = 80\%$<br>$sp_{min} = 97\%$ | Interim analysis | Early stopping for efficacy | 1.5% | | NA |
|  |  | Early stopping for futility | 19.2% |  |  |
|  |  | Transition stop | 60.7% | 63.9% |  |
|  | Final analysis | Maintain null hypothesis | 7.1% | 6.1% | 94.7% |
|  |  | Reject null hypothesis | 11.5% | 9.3% | 5.3% |
|  | Overall power |  | 13% | 10.8% | 5.3% |
|  | Power within studies with final analysis |  | 61.8% | 60.4% | 5.3% |
| More liberal criteria<br>$se_{min} = 75\%$<br>$sp_{min} = 90\%$ | Interim analysis | Early stopping for efficacy | 13.7% | | NA |
|  |  | Early stopping for futility | 5.4% |  |  |
|  |  | Transition stop | 41.7% | 48.3% |  |
|  | Final analysis | Maintain null hypothesis | 11.2% | 11.4% | 73.4% |
|  |  | Reject null hypothesis | 28.0% | 21.2% | 26.6% |
|  | Overall power |  | 41.7% | 34.9% | 26.6% |
|  | Power within studies with final analysis |  | 71.4% | 65.0% | 26.6% |

In the FD, which does not incorporate interim analyses, the null hypothesis was rejected in only 5.3% of simulation runs (corresponding to the statistical power) when the stricter diagnostic thresholds (WHO) were applied. Both adaptive designs enabled interim decisions and allowed for early study termination. In 79.9% of ADs simulations and 83.1% of ADp simulations, the studies were stopped early due to futility or infeasible sample size projections. Early stopping due to efficacy occurred in only 1.5% of cases. The null hypothesis was rejected in the final analysis in 11.5% of ADs simulations and 9.3% of ADp simulations. As a result, the overall power increased to 13.0% in ADs and 10.8% in ADp. Conditional power, calculated among studies that reached the final analysis, was 61.8% for ADs and 60.4% for ADp which indicates a substantially improved likelihood of success when initial conditions were met.

Under the more liberal accuracy thresholds, the performance of all designs improved. The fixed design achieved a power of 26.6%, while the adaptive designs reached 41.7% in ADs and 34.9% in ADp. Early termination occurred less frequently under these criteria and resulted in a higher proportion of studies continuing to the final stage. The null hypothesis was rejected in 28.0% of ADs and 21.2% of ADp simulation runs. Conditional power among studies completing both stages increased to 71.4% for ADs and 65.0% for ADp. While these values still fell short of the conventional target of 80%, they revealed substantial gains relative to the stricter scenarios.

When stricter WHO criteria were applied, the mean required sample size exceeded 10,000 participants. For the liberal criteria, the required sample size remained above 2,000. In comparison, the median sample size observed in studies included in the Cochrane review was fewer than 500 participants which underscores a general lack of statistical power in many real-world studies. Under the liberal criteria, recalculated sample sizes in the adaptive designs closely approximated the theoretical requirements up to the 75th percentile. Under the stricter criteria, however, recalculated sample sizes were often substantially lower due to early termination at interim analyses. Several scenarios projected sample size requirements exceeding 40,500 participants. These were deemed infeasible and were consequently stopped early and resulted in a transition stop.

Figures 5 and 6 display relative bias in estimates of sensitivity and specificity. Sensitivity was systematically underestimated across all three designs, with the largest bias observed in ADp and the smallest in FD. In most cases, the bias ranged between −2.5% and −15%, though it reached −25% in scenarios with low prevalence and small sample sizes. The degree of bias depended on both design features and external conditions. Studies initiated around day 40 of the epidemic yielded the lowest bias, while those beginning at day 100 showed the highest. Bias also increased with faster recruitment rates, smaller initial sample sizes, and higher assumed test performance. These patterns likely reflect the impact of ceiling effects and selection bias introduced by interim decisions in adaptive settings where studies with poor early performance are more likely to stop. The direction and magnitude of this bias have important implications for both interpretation and regulatory confidence in early-phase evaluations.

**Figure 5:**
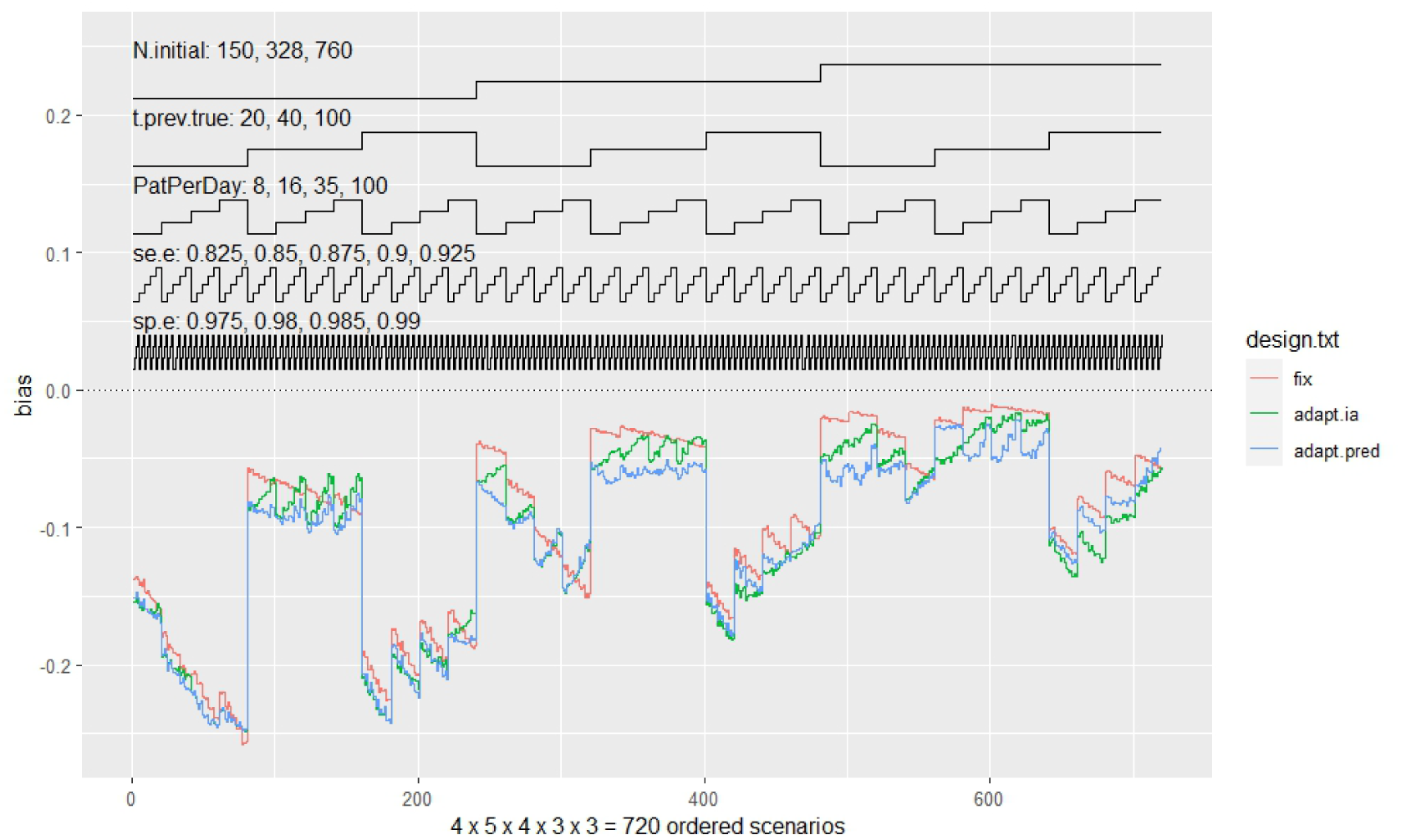
Nested loop plot for the relative bias in sensitivity estimates under WHO criteria.

**Figure 6:**
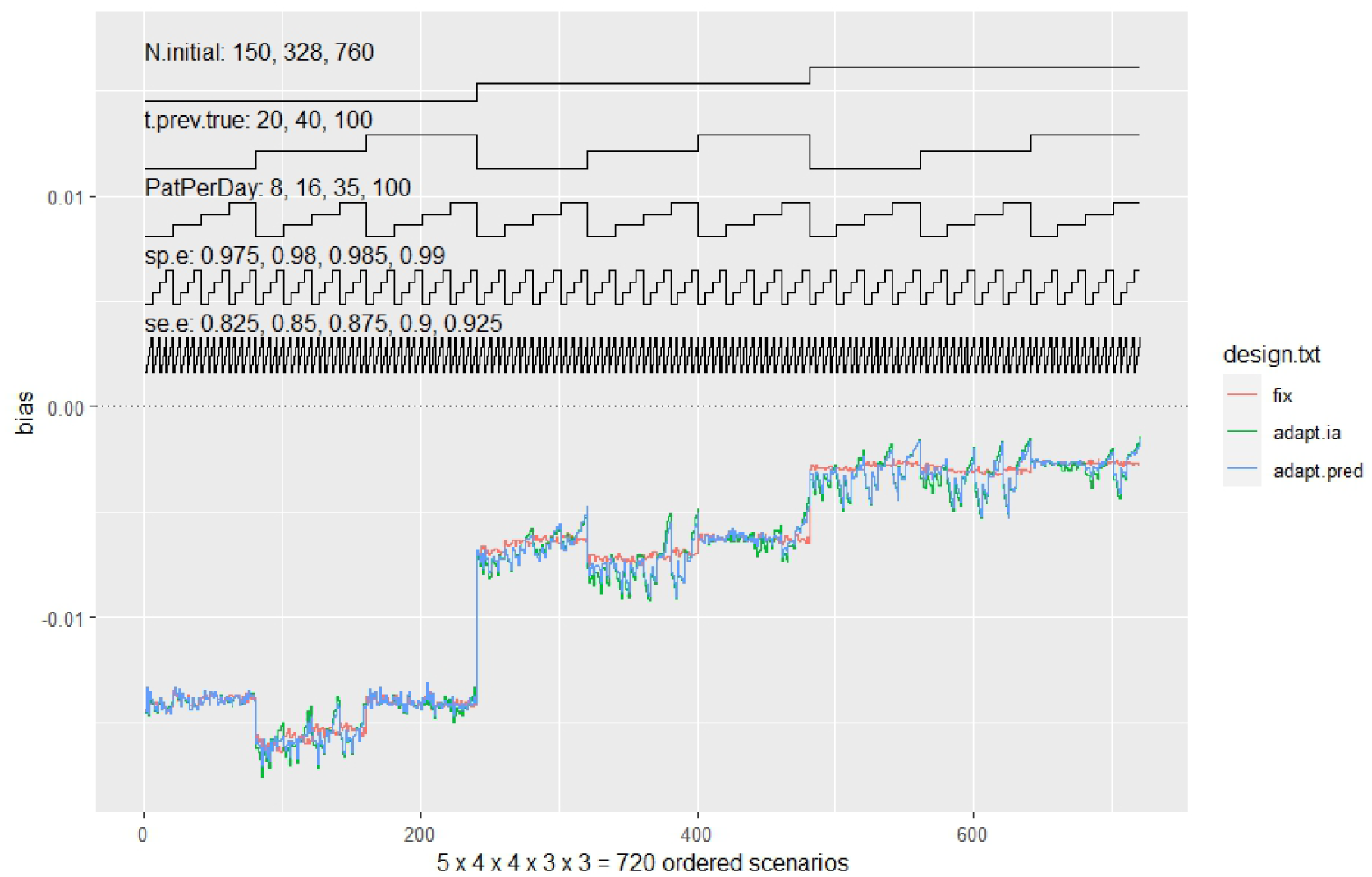
Nested loop plot for the relative bias in specificity estimates under WHO criteria.

Specificity estimates, by contrast, showed minimal bias under all conditions. Figure 6 shows that bias remained below 2% across all designs under the stricter criteria. Under the liberal thresholds the adaptive designs showed slightly less bias than the fixed design (see Appendix). This supports the conclusion that early stopping and sample size adaptation do not compromise specificity estimates in a meaningful way.

With regard to power, the mosaic plot in the Appendix shows that across all scenarios and simulation runs, the total power for all three designs is almost always below 80%. Analogous to the bias, the power increases as the sample size increases, is greatest at 40 days after the start of the pandemic followed by 20 days and is greater for the liberal criteria than for the WHO criteria. Furthermore, the advantage of the adaptive designs is shown here by a greater power than in the fixed design (especially for small and medium initial sample sizes), whereby the ADs design has a greater power than the ADp design. However, the ADs design may result in overpowering, which practically does not occur with the ADp design.

Figure 7 summarizes the power estimates from simulations that reached final analysis. In the FD, power approached 80% only in scenarios characterized by early recruitment, high incidence, and large initial sample sizes. The ADp design achieved adequate power in many scenarios, although not consistently in low-prevalence or small-sample settings. ADs achieved higher power overall but also led to some cases of overpowering. These results highlight the trade-off between power optimization and resource efficiency. ADp may be more conservative but avoids excessive sample size escalation and makes it more suitable in situations where avoiding overpowering is a concern. Taken together, these findings reveal that adaptive designs can increase the efficiency and responsiveness of diagnostic accuracy studies when aligned with epidemic dynamics. However, the benefits depend on careful calibration of decision rules and realistic consideration of operational constraints.

**Figure 7.**
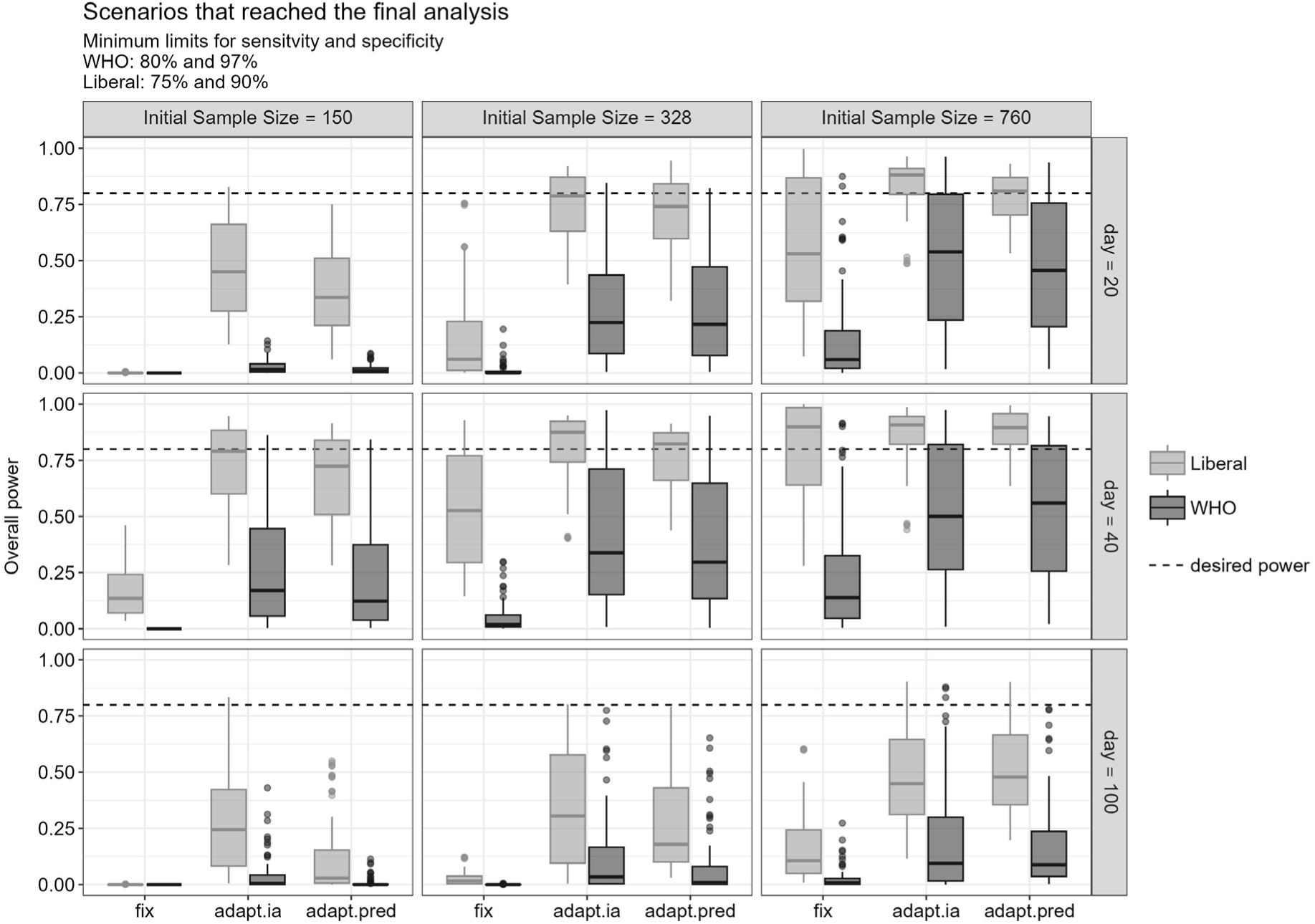
Mosaic plot for the power among simulation runs that reached the final analysis, by design and scenario characteristics.

## 5 Discussion

Diagnostic test accuracy studies conducted during rapidly evolving epidemics with emerging pathogens face several obstacles: unknown prevalence trajectories, sparse early data, and urgency that constrains study duration and achievable sample sizes. The results of this work suggest that adaptive designs may increase operational efficiency under such conditions by enabling interim decisions, sample size adaptation, and preventing the continuation of studies that are unlikely to reach confirmatory conclusions. Compared with a fixed design, both the standard adaptive design and the adaptive design with prevalence prediction allowed early termination of unpromising trials, which reduced the risk of expending large samples on scenarios that offered little chance of meaningful inference. This pattern aligns with prior work on the utility of adaptive features in infectious disease contexts [11, 12] and with arguments for aligning DTA evaluation with epidemic disease models [4].

The standard adaptive design and the adaptive design with prevalence prediction model did not behave identically across scenarios. In WHO-threshold settings, the probability of early termination for futility or infeasibility was high in both designs (with the standard adaptive design being terminated early slightly more often), whereas under more liberal thresholds both designs more frequently completed recruitment and reached a final decision. The prevalence-informed design showed advantages in settings where the projected prevalence trend shifted the expected information gain across recruitment time, enabling more realistic sample size projections under epidemic non-stationarity. These differences suggest a mechanistic trade-off: prevalence projection introduces model-based information that may avert overpowering in scenarios with declining prevalence, while the standard adaptive approach may react faster when early conditional error information strongly disfavours continuation.

Methodologically, these findings support the view that the interaction between recruitment pace, interim stopping, prevalence evolution and decision boundaries is not additive but mutually shaping. The observed bias patterns illustrate this coupling: sensitivity estimates tended to be underestimated more strongly in adaptive settings, while specificity was comparatively robust. This underestimation likely arises from selection effects induced by interim stopping rules: designs stop earlier when interim performance is poor, leaving a non-representative subset of studies to reach final analysis. Such a mechanism is consistent with known properties of curtailed sampling and conditional continuation [8], and should be anticipated when interpreting sensitivity estimates from early-phase DTA studies under adaptive control.

The fact that sample sizes required to meet WHO thresholds exceeded the magnitude typically seen in real-world pandemic studies supports the hypothesis that many existing evaluations were statistically underpowered relative to regulatory benchmarks [2, 3]. Adaptive designs did not solve this constraint in absolute terms, but they made the constraint visible earlier, which is materially different in practice. Declaring infeasibility early is not a statistical failure but a resource-preserving property when target thresholds are structurally unattainable under realistic recruitment. This aligns with prior calls to embed feasibility checks into confirmatory design rather than discover infeasibility post-hoc [6].

Importantly, the interpretation of superiority between the standard adaptive design and the adaptive design with prevalence prediction cannot be collapsed to a single ranking: their relative behaviour differed by threshold regime, timing of study initiation, and prevalence evolution. The trade-off suggests that adaptive designs should not be treated as exchangeable but as design families whose performance depends on alignment between decision rules and epidemic information structure. Under conditions of sharply dynamic prevalence, using a prevalence projection model appears beneficial, whereas under stable or weakly varying prevalence the added model layer may offer little incremental gain. This context-dependence is consistent with the notion that epidemic-sensitive design is not a single method but a design class parameterized by expected disease dynamics [4].

Limitations include reliance on simulation scenarios and assumed structures of epidemic progression that may not fully capture operational variability in real-world deployments. Moreover, the analyses focused on early-phase properties; long-run feedback effects between test adoption and epidemic dynamics were not modelled. Nonetheless, the patterns observed here are coherent across wide factorial conditions and align with principled expectations from adaptive methodology. Taken together, the results indicate that adaptive DTA designs can reduce wasted effort, reveal infeasibility earlier, and modulate sample size decisions in a way that reflects epidemic dynamics without compromising the logical integrity of confirmatory evaluation.

## 6 Conclusion

Adaptive designs appear to improve the operational efficiency of diagnostic accuracy studies during epidemics by enabling earlier detection of infeasible or non-promising scenarios and by aligning sample size decisions more closely with epidemic dynamics. The standard adaptive design and the adaptive design with prevalence prediction model exhibited context-dependent advantages rather than a clear ranking. The results suggest that confirmatory DTA studies for emerging infections may benefit from adaptive planning when epidemic dynamics generate unstable information conditions.

## Data Availability

All data produced in the present study are available upon reasonable request to the authors

## Appendix

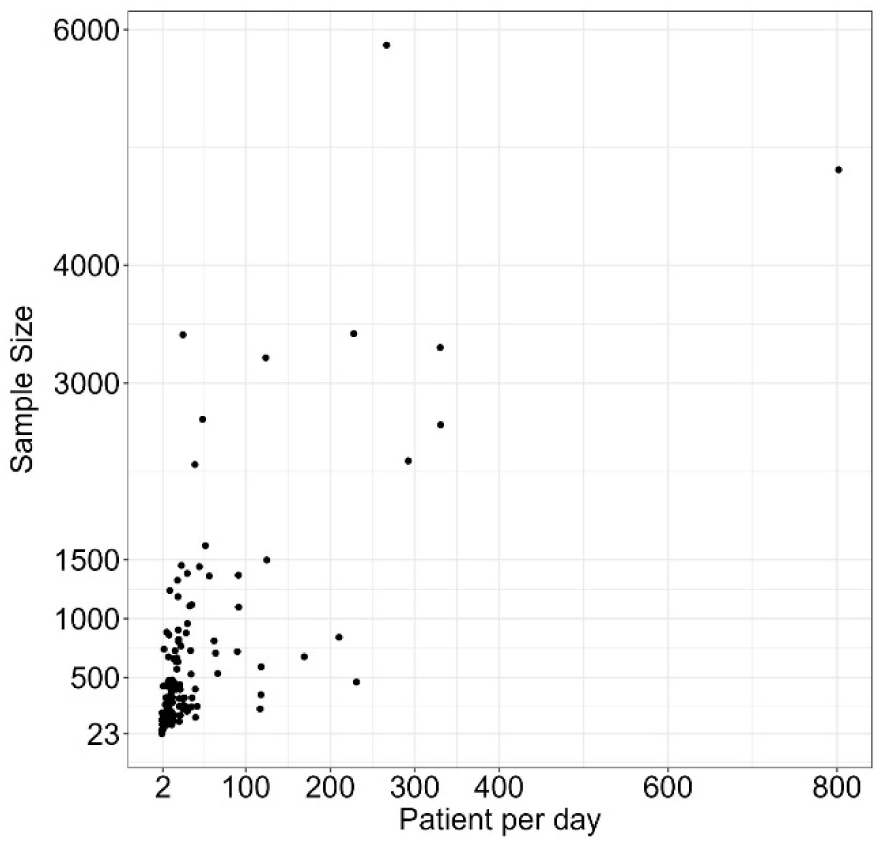
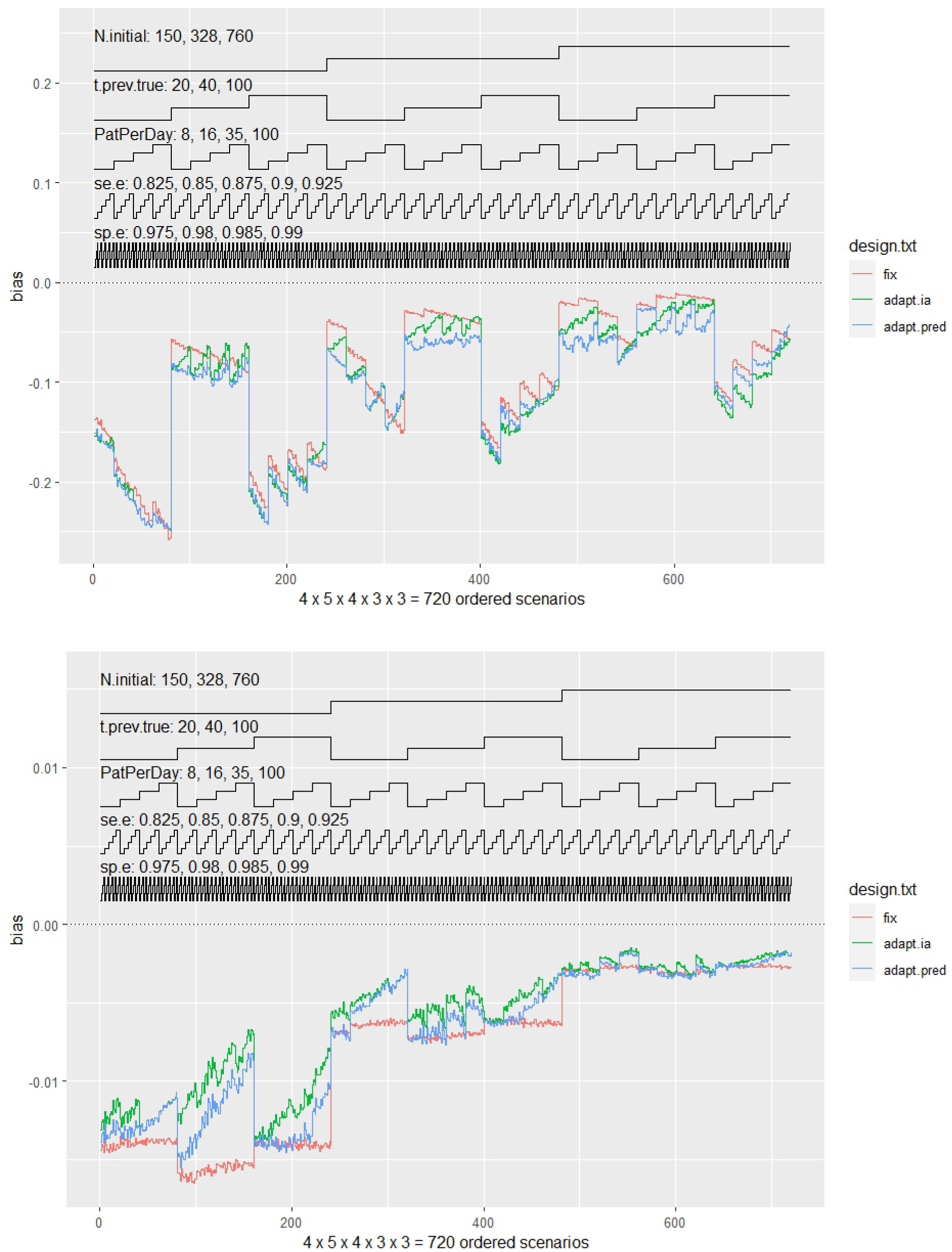
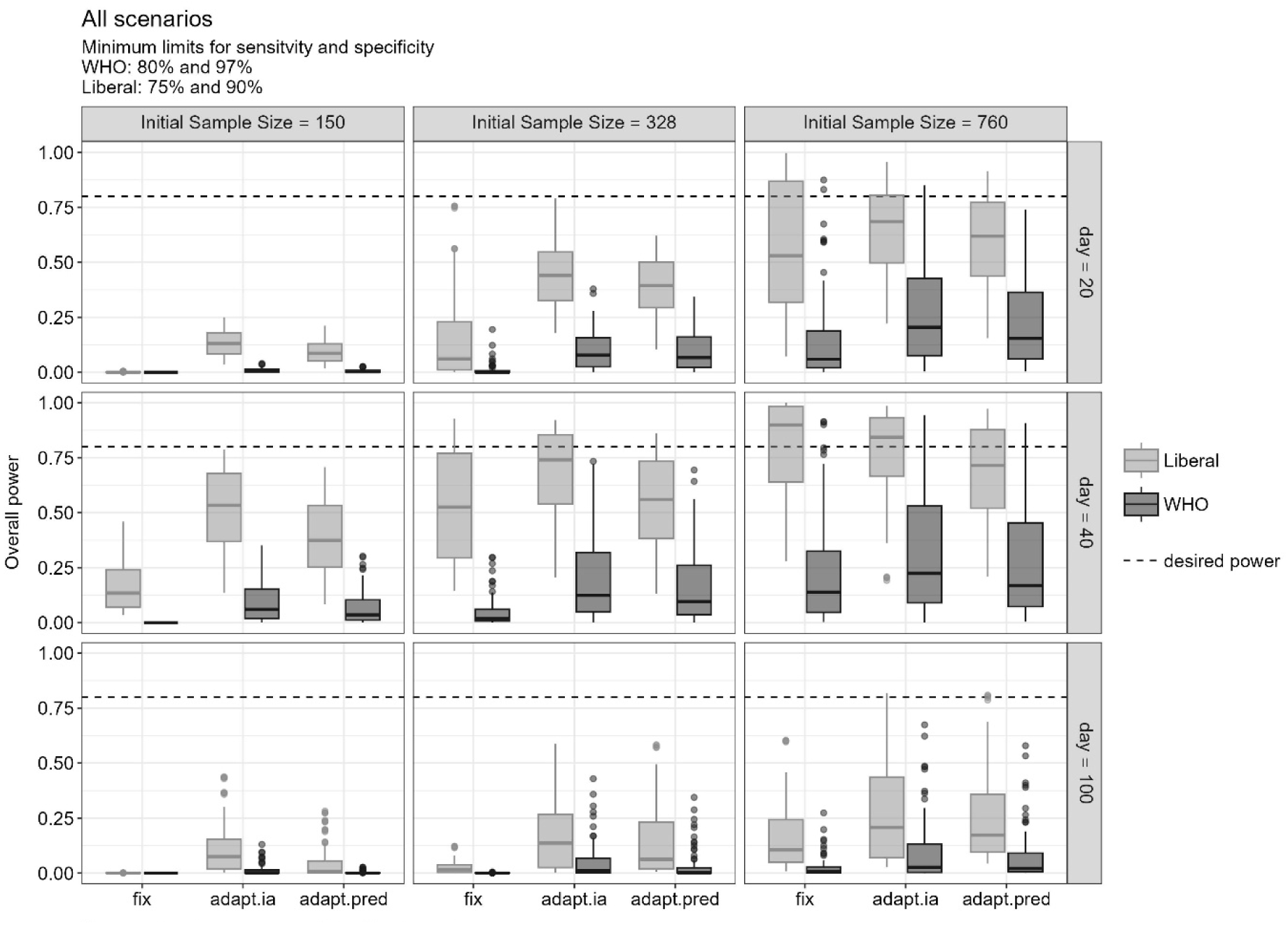

## References

1. WHO, Target product profiles for priority diagnostics to support response to the COVID-19 pandemic v. 1.0. 2020.

2. WHO. Antigen-detection in the diagnosis of SARS-CoV-2 infection: Interim guidance. 2021; Available from: https://www.who.int/publications/i/item/antigen-detection-in-the-diagnosis-of-sars-cov-2infection-using-rapid-immunoassays.

3. Dinnes, J., et al., Rapid, point-of-care antigen tests for diagnosis of SARS-CoV-2 infection. Cochrane database of systematic reviews, 2022(7).

4. Chaturvedi, M., et al., The impact of inaccurate assumptions about antibody test accuracy on the parametrisation and results of infectious disease models of epidemics. Epidemics, 2024. 46: p. 100741.

5. Chaturvedi, M., et al., Corrigendum to “The impact of inaccurate assumptions about antibody test accuracy on the parametrisation and results of infectious disease models of epidemics”[Epidemics 46 (2024) 100741]. Epidemics, 2024. 47: p. 100766.

6. Korevaar, D.A., et al., Targeted test evaluation: a framework for designing diagnostic accuracy studies with clear study hypotheses. Diagnostic and prognostic research, 2019. 3(1): p. 22.

7. Obuchowski, N.A., Sample size calculations in studies of test accuracy. Statistical Methods in Medical Research, 1998. 7(4): p. 371–392.

8. Stark, M. and A. Zapf, Sample size calculation and re-estimation based on the prevalence in a single-arm confirmatory diagnostic accuracy study. Statistical Methods in Medical Research, 2020. 29(10): p. 2958–2971.

9. Morris, T.P., I.R. White, and M.J. Crowther, Using simulation studies to evaluate statistical methods. Statistics in medicine, 2019. 38(11): p. 2074–2102.

10. Burton, A., et al., The design of simulation studies in medical statistics. Statistics in medicine, 2006. 25(24): p. 4279–4292.

11. Bauer, P. and K. Köhne, Evaluation of experiments with adaptive interim analyses. Biometrics, 1994: p. 1029–1041.

12. Proschan, M.A. and S.A. Hunsberger, Designed extension of studies based on conditional power. Biometrics, 1995: p. 1315–1324.

